# On-site AASM-accredited sleep facility status and hospitalization outcomes among adults admitted with acute hypercapnic respiratory failure: a retrospective cohort study

**DOI:** 10.64898/2026.07.28.26359158

**Authors:** Alec R. Flores, Saad Bin Jamil, Gregory D. Gudleski, Alberto F. Monegro

## Abstract

**Objectives:** Acute hypercapnic respiratory failure (AHRF) is a common cause of hospitalization among adults with chronic respiratory and sleep-related disorders. We evaluated whether admission to hospitals with an on-site American Academy of Sleep Medicine (AASM)-accredited sleep facility was associated with length of stay (LOS) and hospitalization cost among adults admitted with AHRF.

**Methods:** We conducted a retrospective cohort study using the Healthcare Cost and Utilization Project New York State Inpatient Database from 2017 to 2021. Adults admitted to hospitals outside New York City were included because hospital density, referral patterns, and access to specialty services differ from those in New York City. Hospitals were classified by the presence of an on-site AASM-accredited sleep facility, used as a structural proxy for institutional sleep medicine capacity. LOS and hospitalization cost were log10-transformed before analysis. We used bivariate analyses and hierarchical multivariable linear regression models adjusting for demographic, clinical, socioeconomic, and hospital-level covariates.

**Results:** A total of 3,247 adults met inclusion criteria. In unadjusted analyses, admission to hospitals with an on-site AASM-accredited sleep facility was associated with shorter LOS and lower hospitalization cost. After adjustment, sleep facility status was not independently associated with either outcome and did not improve model fit. Greater comorbidity burden was the strongest independent predictor of both longer LOS and higher cost. Housing instability and race categorized as other than White were also associated with higher resource use. Chronic obstructive pulmonary disease was independently associated with shorter LOS and lower cost.

**Conclusions:** Among adults hospitalized with AHRF, LOS and hospitalization cost were more strongly associated with comorbidity burden and socioeconomic disadvantage than with the presence of an on-site AASM-accredited sleep facility. These findings suggest that accreditation status alone may not capture inpatient sleep medicine processes most relevant to acute respiratory care.

## Introduction

Sleep-related disorders contribute substantially to healthcare utilization, costs, and morbidity, with effects extending beyond outpatient management into acute care settings. In the United States, sleep disorders account for a substantial share of healthcare expenditures.^1^ Among these, sleep-disordered breathing is strongly associated with cardiovascular, metabolic, and respiratory complications across diverse patient populations.^2–4^ These associations are particularly relevant in patients with chronic ventilatory disorders, obesity hypoventilation syndrome, and related conditions that may culminate in acute hypercapnic respiratory failure (AHRF).

AHRF is a frequent and clinically important cause of hospitalization, particularly among adults with chronic respiratory disease and sleep-related breathing disorders. These admissions are often associated with prolonged length of stay, substantial resource utilization, and recurrent hospital use. Because AHRF often reflects a mixture of pulmonary disease severity, nocturnal hypoventilation, and sleep-disordered breathing, sleep medicine expertise may plausibly influence care. However, the role of sleep medicine during hospitalization may vary substantially across institutions. Inpatient management of AHRF is typically led by pulmonary, critical care, and respiratory therapy teams using established protocols for ventilatory stabilization and noninvasive ventilation.^5-6^ As a result, the presence of outpatient sleep medicine resources may not necessarily translate into measurable differences in acute inpatient outcomes.

More than 2,800 sleep facilities in the United States are accredited by the American Academy of Sleep Medicine (AASM).^7^ The presence of an on-site accredited sleep facility may reflect broader institutional capacity for sleep-related services, including diagnostic testing, specialty expertise, and continuity with outpatient sleep care. However, AASM accreditation does not necessarily imply routine inpatient consultation, inpatient polysomnography, standardized management pathways for hospitalized patients, or integrated transitional care after discharge. Accordingly, on-site AASM-accredited sleep facility status may serve as a structural marker of sleep medicine infrastructure rather than a direct measure of inpatient sleep service availability.

This distinction is important because outcomes among patients hospitalized with AHRF are shaped not only by clinical severity, but also by comorbidity burden and social determinants of health. Socioeconomic disadvantage and related structural factors are consistently associated with healthcare utilization, resource use, and costs.^8-9^ In patients with sleep-related and chronic ventilatory disorders, these factors may influence timely diagnosis, access to outpatient sleep evaluation, treatment adherence, and the ability to coordinate follow-up after hospitalization. Thus, structural hospital resources may be insufficient to offset the effects of underlying clinical and social complexity.

We therefore evaluated whether admission to hospitals with on-site AASM-accredited sleep facilities was associated with differences in length of stay and hospitalization cost among adults admitted with AHRF. We hypothesized that hospitals with accredited sleep facilities might demonstrate differences in hospitalization outcomes reflecting broader institutional resources and specialty infrastructure, but we also recognized that accreditation status alone may not reflect the presence of inpatient sleep medicine consultation services, integrated ventilatory care pathways, or transitional sleep health support. By examining both hospital infrastructure and patient-level social and clinical factors, this study seeks to clarify whether sleep medicine capacity at the hospital level is associated with acute outcomes in a condition closely linked to sleep-related respiratory disease.

## Methods

### Study design

This retrospective cohort study included adults admitted to New York State hospitals between January 1, 2017, and December 31, 2021, with a primary diagnosis code consistent with acute hypercapnic respiratory failure or obesity hypoventilation-related illness (J96.02, J96.22, E66.20). This study involved human participants and was exempted by the University at Buffalo Institutional Review Board (FWA00008824).

### Participants

Patients younger than 18 years, admitted outside New York State or to hospitals located in New York City, transferred during hospitalization, died before discharge, or were admitted to hospitals lacking a National Provider Identifier were excluded to ensure complete and interpretable length-of-stay and hospitalization cost data **(Figure 1)**.

**Figure 1.**
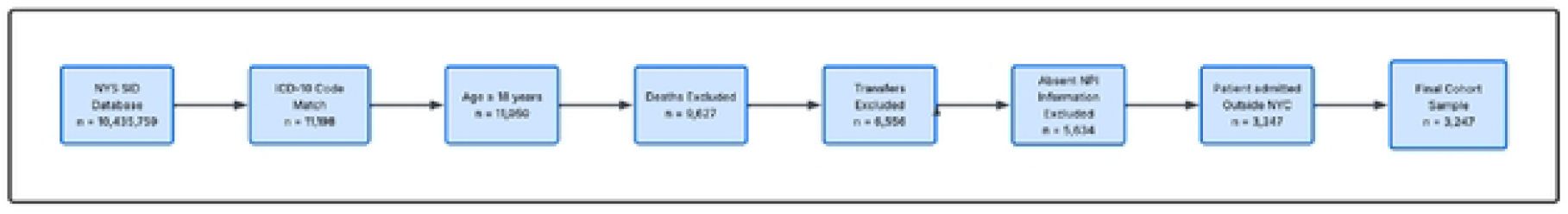
STROBE flow diagram of study selection.

### Data source

Data were obtained from the Healthcare Cost and Utilization Project (HCUP) New York State Inpatient Database and first accessed on July 3, 2024, for research purposes. The dataset was de-identified, and the authors did not have access to information that could identify individual participants during or after data collection. Demographic variables included age, sex, race, and housing status. Comorbid conditions, including hypertension, congestive heart failure, diabetes mellitus, chronic obstructive pulmonary disease, obstructive sleep apnea, and end-stage renal disease, were identified using ICD-10 billing codes and treated as binary indicators. Comorbidity burden was operationalized as the total number of chronic conditions documented in the administrative record. The presence of an on-site AASM-accredited sleep facility was verified using the AASM online directory and used as a structural proxy for institutional sleep medicine capacity.

### Statistical analysis

Descriptive statistics were used to summarize demographic and clinical variables for the overall sample. Length of stay and hospitalization cost were right-skewed and therefore log10-transformed before analysis; the transformed variables approximated normal distributions. We used Welch t tests for two-group comparisons and Pearson correlations for continuous variables to examine unadjusted associations with the log-transformed outcomes. Hierarchical linear regression models were then used to identify independent predictors of log-transformed length of stay and hospitalization cost, with on-site AASM-accredited sleep facility status entered in the final step. Because the analysis was exploratory and the dataset included a limited number of variables with sufficient case counts, all demographic and clinical covariates were retained in the models. Statistical analyses were conducted using SPSS version 29, with a significance threshold of p < .05.

### Patient and public involvement

It was not appropriate or possible to involve patients or the public in the design, conduct, reporting, or dissemination plans of our research.

## Results

### Study participants

A total of 3,247 adults met inclusion criteria **(Table 1)**. The mean age was 64.5 years; most patients identified as White (80.5%) and female (54.8%). The majority were admitted to hospitals without on-site AASM-accredited sleep facilities (91.0%). Multimorbidity was prominent, with participants averaging 17.6 chronic conditions documented in the HCUP database. Chronic obstructive pulmonary disease was the most prevalent comorbidity (77.1%), followed by congestive heart failure (48.0%), diabetes mellitus (38.6%), hypertension (27.4%), and obstructive sleep apnea (21.7%).

**Table 1.**
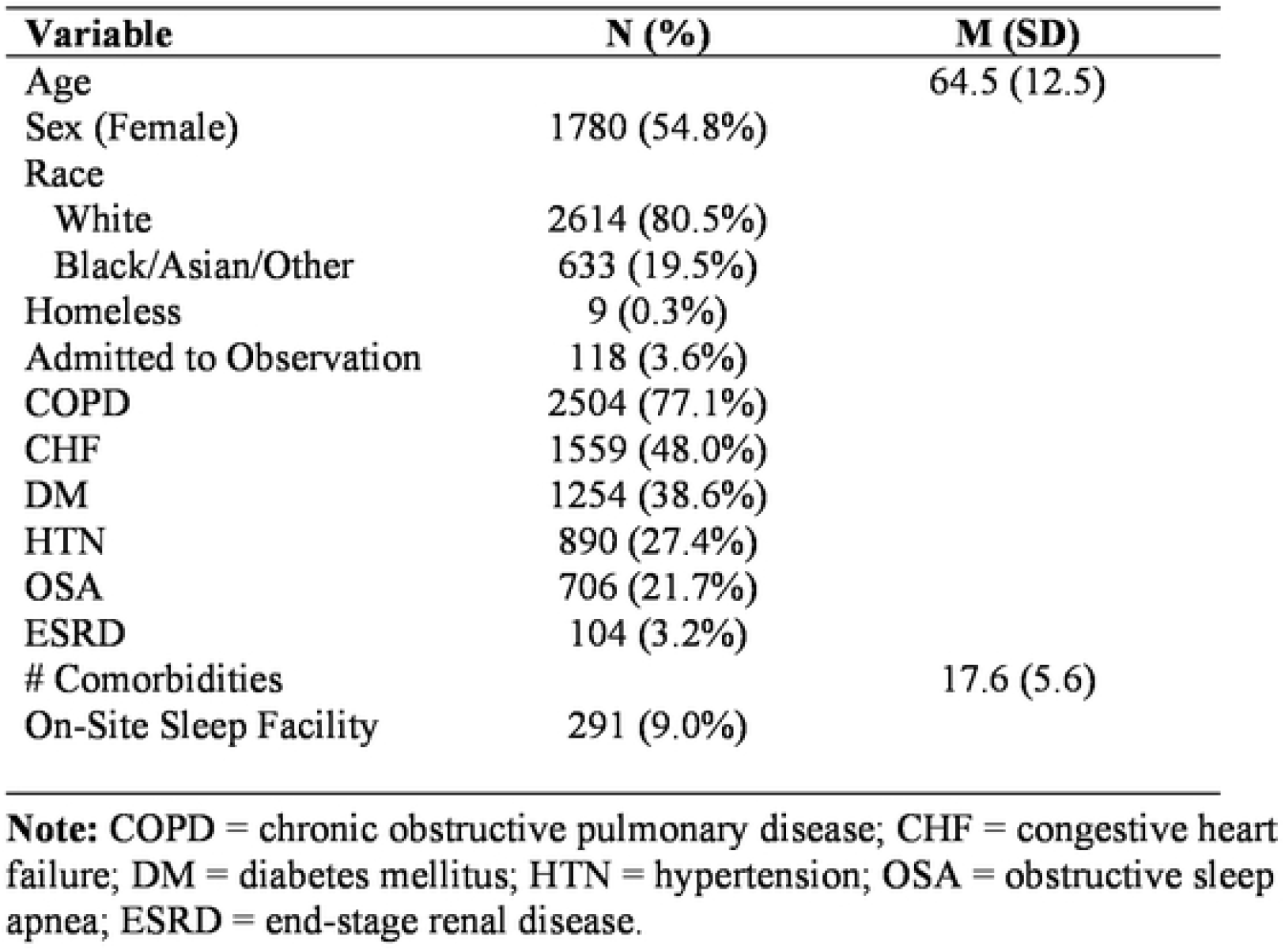
Baseline characteristics of the study sample (N = 3247).

| **Variable** | **N (%)** | **M (SD)** |
| --- | --- | --- |
| Age |  | 64.5 (12.5) |
| Sex (Female) | 1780 (54.8%) |  |
| Race |  |  |
| White | 2614 (80.5%) |  |
| Black/Asian/Other | 633 (19.5%) |  |
| Homeless | 9 (0.3%) |  |
| Admitted to Observation | 118 (3.6%) |  |
| COPD | 2504 (77.1%) |  |
| CHF | 1559 (48.0%) |  |
| DM | 1254 (38.6%) |  |
| HTN | 890 (27.4%) |  |
| OSA | 706 (21.7%) |  |
| ESRD | 104 (3.2%) |  |
| # Comorbidities |  | 17.6 (5.6) |
| On-Site Sleep Facility | 291 (9.0%) |  |

**Note:** COPD = chronic obstructive pulmonary disease; CHF = congestive heart failure; DM = diabetes mellitus; HTN = hypertension; OSA = obstructive sleep apnea; ESRD = end-stage renal disease.

### Length of stay

In bivariate analyses, longer log-transformed length of stay was associated with older age, homelessness, observation status, congestive heart failure, and higher comorbidity burden. Shorter log-transformed length of stay was observed among patients with chronic obstructive pulmonary disease or hypertension and among those admitted to hospitals with on-site AASM-accredited sleep facilities **(Table 2)**. In hierarchical regression modeling, covariates entered in Step 1 accounted for 14.5% of the variance in log-transformed length of stay (p < .001). Addition of on-site AASM-accredited sleep facility status in Step 2 did not significantly improve model fit (ΔR^2^ = .001, p = .697). Greater comorbidity burden was the strongest independent predictor of log-transformed length of stay (β = .377, p < .001), followed by chronic obstructive pulmonary disease (β = −.078, p < .001) and homelessness (β = .042, p = .009) **(Table 3 and Figure 2)**.

**Table 2.**
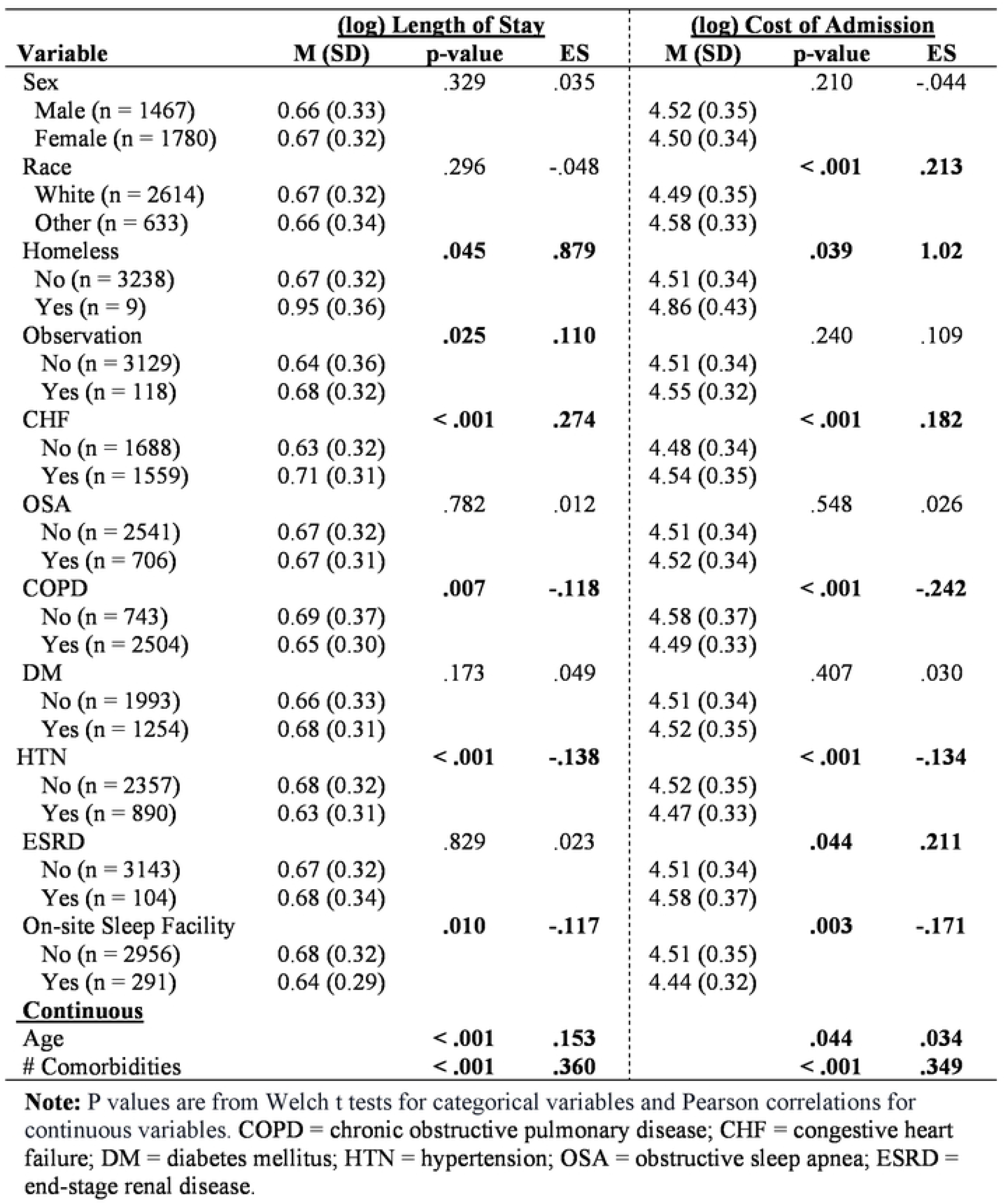
Bivariate analyses of clinical characteristics and log-transformed outcomes (N = 3247).

| Variable | <u>(log) Length of Stay</u> |  |  | <u>(log) Cost of Admission</u> |  |  |
| --- | --- | --- | --- | --- | --- | --- |
|  | M (SD) | p-value | ES | M (SD) | p-value | ES |
| Sex |  | .329 | .035 |  | .210 | -.044 |
| Male (n = 1467) | 0.66 (0.33) |  |  | 4.52 (0.35) |  |  |
| Female (n = 1780) | 0.67 (0.32) |  |  | 4.50 (0.34) |  |  |
| Race |  | .296 | -.048 |  | < .001 | .213 |
| White (n = 2614) | 0.67 (0.32) |  |  | 4.49 (0.35) |  |  |
| Other (n = 633) | 0.66 (0.34) |  |  | 4.58 (0.33) |  |  |
| Homeless |  | .045 | .879 |  | .039 | 1.02 |
| No (n = 3238) | 0.67 (0.32) |  |  | 4.51 (0.34) |  |  |
| Yes (n = 9) | 0.95 (0.36) |  |  | 4.86 (0.43) |  |  |
| Observation |  | .025 | .110 |  | .240 | .109 |
| No (n = 3129) | 0.64 (0.36) |  |  | 4.51 (0.34) |  |  |
| Yes (n = 118) | 0.68 (0.32) |  |  | 4.55 (0.32) |  |  |
| CHF |  | < .001 | .274 |  | < .001 | .182 |
| No (n = 1688) | 0.63 (0.32) |  |  | 4.48 (0.34) |  |  |
| Yes (n = 1559) | 0.71 (0.31) |  |  | 4.54 (0.35) |  |  |
| OSA |  | .782 | .012 |  | .548 | .026 |
| No (n = 2541) | 0.67 (0.32) |  |  | 4.51 (0.34) |  |  |
| Yes (n = 706) | 0.67 (0.31) |  |  | 4.52 (0.34) |  |  |
| COPD |  | .007 | -.118 |  | < .001 | -.242 |
| No (n = 743) | 0.69 (0.37) |  |  | 4.58 (0.37) |  |  |
| Yes (n = 2504) | 0.65 (0.30) |  |  | 4.49 (0.33) |  |  |
| DM |  | .173 | .049 |  | .407 | .030 |
| No (n = 1993) | 0.66 (0.33) |  |  | 4.51 (0.34) |  |  |
| Yes (n = 1254) | 0.68 (0.31) |  |  | 4.52 (0.35) |  |  |
| HTN |  | < .001 | -.138 |  | < .001 | -.134 |
| No (n = 2357) | 0.68 (0.32) |  |  | 4.52 (0.35) |  |  |
| Yes (n = 890) | 0.63 (0.31) |  |  | 4.47 (0.33) |  |  |
| ESRD |  | .829 | .023 |  | .044 | .211 |
| No (n = 3143) | 0.67 (0.32) |  |  | 4.51 (0.34) |  |  |
| Yes (n = 104) | 0.68 (0.34) |  |  | 4.58 (0.37) |  |  |
| On-site Sleep Facility |  | .010 | -.117 |  | .003 | -.171 |
| No (n = 2956) | 0.68 (0.32) |  |  | 4.51 (0.35) |  |  |
| Yes (n = 291) | 0.64 (0.29) |  |  | 4.44 (0.32) |  |  |
| **<u>Continuous</u>** |  |  |  |  |  |  |
| Age |  | < .001 | .153 |  | .044 | .034 |
| # Comorbidities |  | < .001 | .360 |  | < .001 | .349 |

**Note:** P values are from Welch t tests for categorical variables and Pearson correlations for continuous variables. COPD = chronic obstructive pulmonary disease; CHF = congestive heart failure; DM = diabetes mellitus; HTN = hypertension; OSA = obstructive sleep apnea; ESRD = end-stage renal disease.

**Table 3.** Hierarchical linear regression predicting log-transformed length of stay (N = 3247).

| Variable | <u>Step 1</u> |  |  |  | <u>Step 2</u> |  |  |  |
| --- | --- | --- | --- | --- | --- | --- | --- | --- |
| | B | 95% CI | $\beta$ | p-value | B | 95% CI | $\beta$ | p-value |
| Age | .001 | [.000, .002] | .039 | .023 | .001 | [.000, .002] | .039 | .024 |
| Sex | .010 | [-.011, .031] | .016 | .338 | .010 | [-.011, .031] | .015 | .346 |
| Race | -.001 | [-.011, .009] | -.003 | .854 | -.001 | [-.011, .009] | -.003 | .865 |
| Homeless | .258 | [.063, .454] | .042 | .010 | .259 | [.063, .454] | .042 | .009 |
| Observation | -.057 | [-.112, -.002] | -.033 | .042 | -.057 | [-.112, -.002] | -.033 | .041 |
| COPD | -.051 | [-.074, -.029] | -.078 | < .001 | -.052 | [-.074, -.029] | -.078 | < .001 |
| CHF | .017 | [-.009, .044] | .027 | .206 | .017 | [-.010, .044] | .027 | .211 |
| DM | -.023 | [-.048, .002] | -.030 | .072 | -.023 | [-.048, .002] | -.030 | .070 |
| HTN | -.008 | [-.036, .020] | -.011 | .581 | -.008 | [-.036, .020] | -.011 | .576 |
| OSA | -.034 | [-.060, -.008] | -.043 | .009 | -.034 | [-.060, -.008] | -.044 | .009 |
| ESRD | -.045 | [-.104, .014] | -.025 | .134 | -.045 | [-.104, .014] | -.025 | .133 |
| # Comorbidities | .022 | [.020, .024] | .377 | < .001 | .022 | [.020, .024] | .378 | < .001 |
| On-Site Sleep Facility | - | - | - | - | .007 | [-.029, .043] | .006 | .697 |
| Adjusted $R^2$ | | .145 | | < .001 | | .146 | | < .001 |
| Adjusted $\Delta R^2$ | | - | | - | | .001 | | .697 |

**Note:** COPD = chronic obstructive pulmonary disease; CHF = congestive heart failure; DM = diabetes mellitus; HTN = hypertension; OSA = obstructive sleep apnea; ESRD = end-stage renal disease.

**Figure 2.**
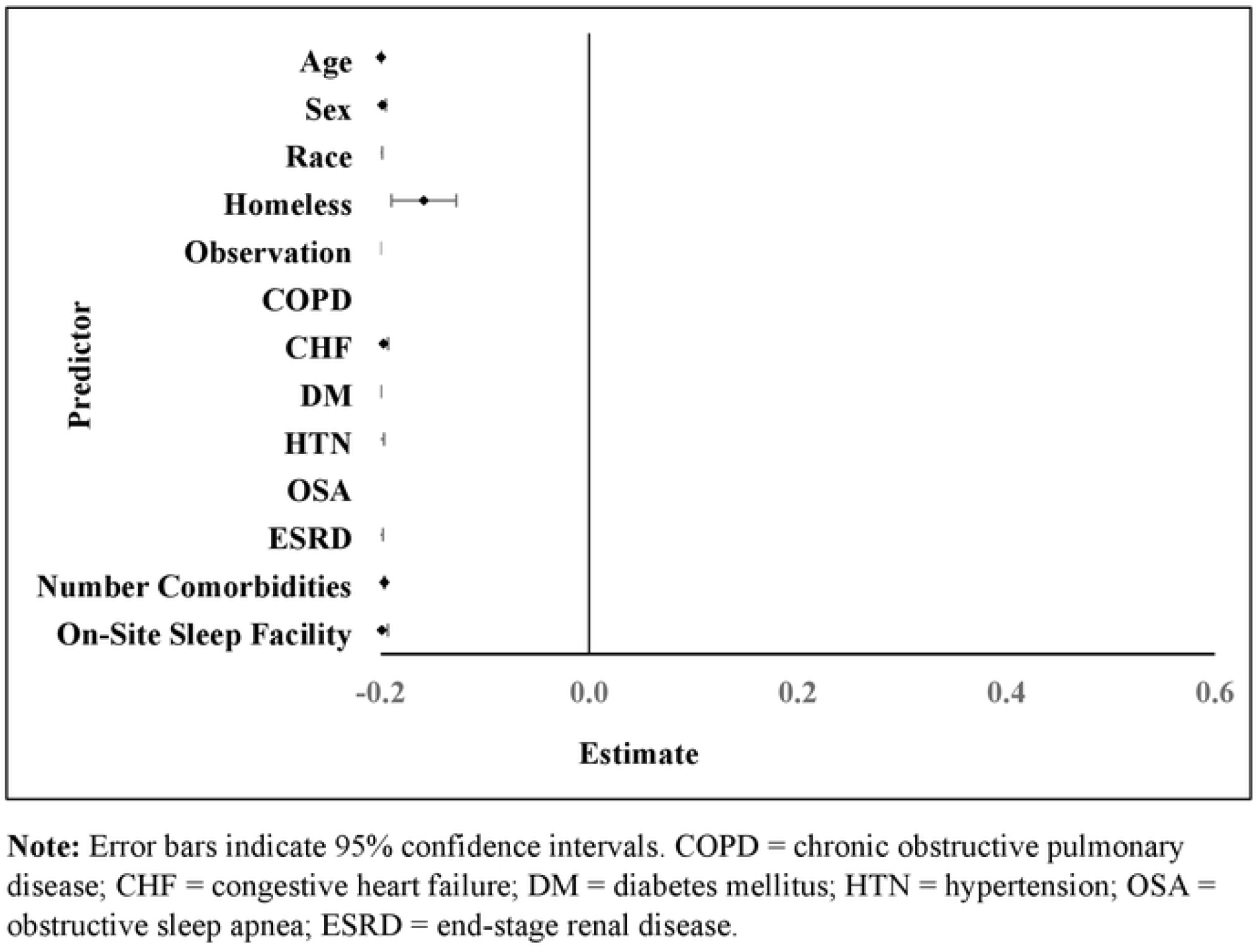
Coefficient estimates for variables in final regression model predicting length of stay.

### Hospitalization cost

Patterns observed for log-transformed hospitalization cost were similar to those for length of stay. Higher log-transformed hospitalization cost was associated with homelessness, congestive heart failure, end-stage renal disease, higher comorbidity burden, and race categorized as other than White. Lower log-transformed hospitalization cost was observed among younger patients, those with chronic obstructive pulmonary disease or hypertension, and those admitted to hospitals with on-site AASM-accredited sleep facilities. The final hierarchical regression model explained 16.2% of the variability in log-transformed hospitalization cost (p < .001).

Comorbidity burden was again the strongest independent predictor (β = .399, p < .001), followed by chronic obstructive pulmonary disease (β = −.093, p < .001), race categorized as other than White (β = .086, p < .001), diabetes mellitus (β = −.091, p < .001), age (β = −.065, p = .001), homelessness (β = .052, p = .001), and obstructive sleep apnea (β = −.039, p = .019). On-site AASM-accredited sleep facility status was not independently associated with log-transformed hospitalization cost after adjustment (ΔR^2^ = .001, p = .732) **(Table 4 and Figure 3)**.

**Table 4.** Hierarchical linear regression predicting log transformed hospitalization cost(N= 3247).

| Variable | <u>Step 1</u> |  |  |  | <u>Step 2</u> |  |  |  |
| --- | --- | --- | --- | --- | --- | --- | --- | --- |
| | B | 95% CI | $\beta$ | p-value | B | 95% CI | $\beta$ | p-value |
| Age | -.002 | [-.003, -.001] | -.065 | < .001 | -.002 | [-.003, -.001] | -.065 | < .001 |
| Sex | -.008 | [-.030, .014] | -.012 | .465 | -.008 | [-.030, .014] | -.012 | .466 |
| Race | .028 | [.018, .039] | .089 | < .001 | .028 | [.018, .039] | .086 | < .001 |
| Homeless | .338 | [.131, .546] | .052 | .001 | .338 | [.131, .546] | .052 | .001 |
| Observation | .028 | [-.031, .086] | .015 | .353 | .028 | [-.031, .086] | .015 | .353 |
| COPD | -.076 | [-.102, -.049] | -.093 | < .001 | -.076 | [-.103, -.049] | -.093 | < .001 |
| CHF | -.006 | [-.034, .023] | -.008 | .687 | -.006 | [-.034, .023] | -.008 | .687 |
| DM | -.064 | [-.088, -.040] | -.091 | < .001 | -.064 | [-.088, -.040] | -.091 | < .001 |
| HTN | -.016 | [-.046, .013] | -.021 | .281 | -.016 | [-.046, .013] | -.021 | .281 |
| OSA | -.032 | [-.060, -.005] | -.039 | .019 | -.032 | [-.060, -.005] | -.039 | .019 |
| ESRD | .009 | [-.053, .072] | -.005 | .772 | .009 | [-.053, .072] | -.005 | .772 |
| # Comorbidities | .024 | [.022, .027] | .399 | < .001 | .024 | [.022, .027] | .399 | < .001 |
| On-Site Sleep Facility | - | - | - | - | .006 | [-.030, .039] | .004 | .732 |
| Adjusted $R^2$ | | .161 | | < .001 | | .162 | | < .001 |
| Adjusted $\Delta R^2$ | | - | | - | | .001 | | .732 |

**Note:** COPD = chronic obstructive pulmonary disease; CHF = congestive heart failure; DM = diabetes mellitus; HTN = hypertension; OSA = obstructive sleep apnea; ESRD = end-stage renal disease.

**Figure 3.**
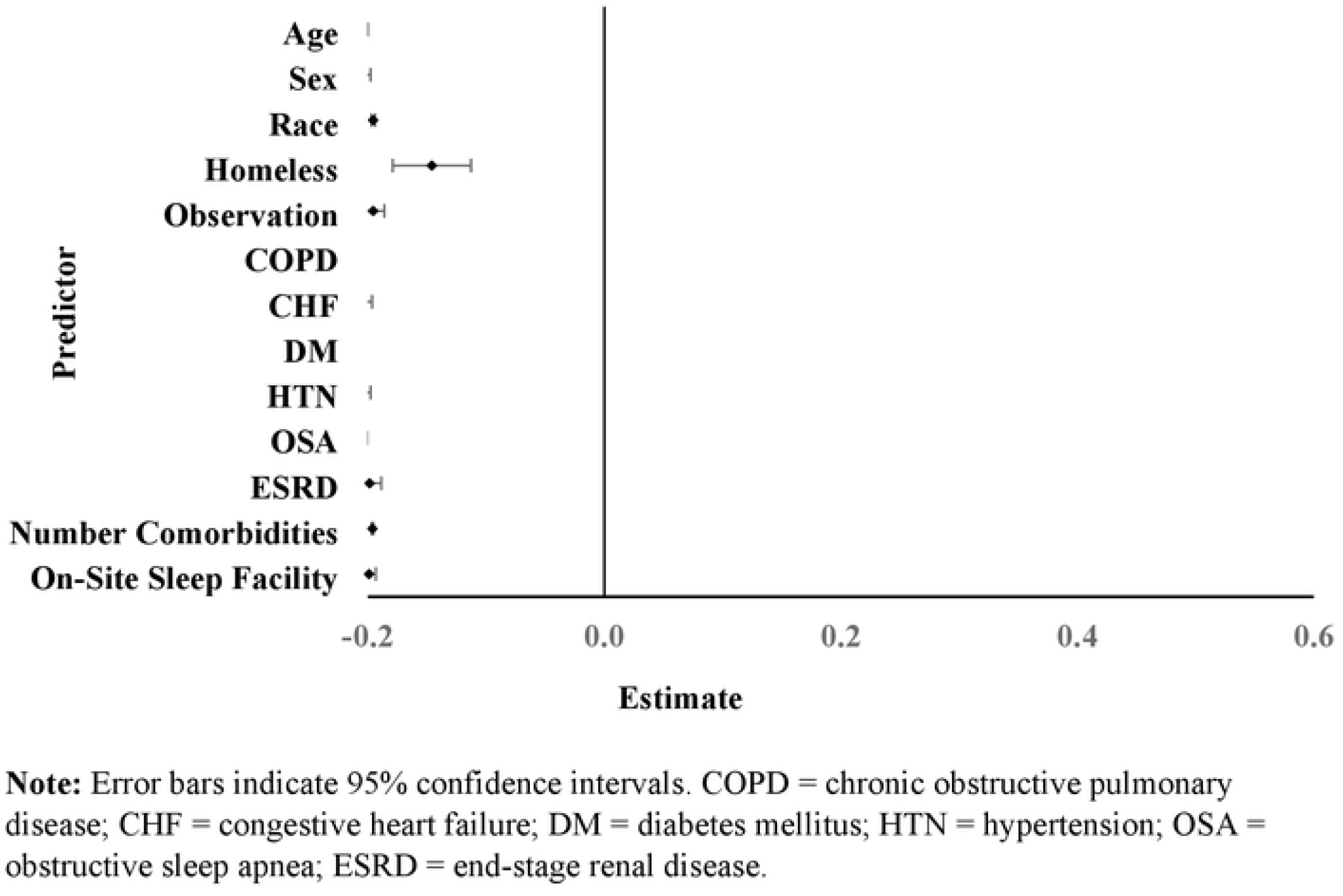
Coefficient estimates for variables in final regression model predicting hospitalization cost.

## Discussion

The principal finding of this study is that on-site AASM-accredited sleep facility status was not independently associated with length of stay or hospitalization cost after adjustment for clinical and socioeconomic factors. Although unadjusted analyses suggested more favorable outcomes at hospitals with accredited sleep facilities, the incremental contribution of this structural marker was negligible after adjustment. These findings suggest that accreditation status alone may be an insufficient surrogate for the inpatient processes of care most relevant to AHRF.

More broadly, the findings underscore the distinction between structural measures of healthcare capacity and actual care delivery processes. Accreditation reflects institutional expertise and resources, but it does not necessarily indicate inpatient consultation availability, protocolized management, or coordinated transitions of care. Thus, accreditation status alone may not capture the mechanisms through which sleep medicine influences acute outcomes.^10-11^

This finding is consistent with the possibility that AHRF is primarily managed through pulmonary, critical care, and respiratory therapy pathways, with sleep medicine services functioning mainly in outpatient settings. In many institutions, accredited sleep facilities may have limited direct involvement in acute respiratory admissions. Accordingly, patients hospitalized with AHRF may receive similar acute management regardless of whether an on-site sleep facility is present.^5-6,12^

Consistent with prior work, socioeconomic and demographic disparities were evident. Race categorized as other than White and homelessness were associated with higher hospitalization costs, underscoring the persistent influence of structural inequities on healthcare utilization and resource use. These findings reinforce the importance of addressing social determinants of health in patients with chronic respiratory and sleep-related disease.^8-9^

Future research should evaluate more direct measures of inpatient sleep medicine involvement, including consultation services, standardized positive airway pressure initiation pathways, multidisciplinary hypoventilation programs, and transitional care models. Outcomes such as readmission, outpatient follow-up, and post-discharge adherence may be more sensitive to the influence of sleep medicine expertise than hospitalization metrics such as length of stay and hospitalization cost.^12-13^

Several limitations should be considered. This retrospective analysis relied on administrative data, which are subject to coding variability and may not fully capture disease severity. AASM accreditation status was used as a proxy for sleep medicine infrastructure but did not measure inpatient sleep consultations, initiation of positive airway pressure therapy, or availability of prior sleep studies. The analysis was limited to AASM-accredited sleep facilities, excluding sleep facilities that may have been accredited by other organizations. Hospitals with AASM-accredited sleep facilities may also differ systematically from those without such facilities in ways not captured in the dataset, including teaching status, subspecialty availability, referral patterns, and other institutional characteristics. Residual confounding from these hospital-level factors may have influenced the observed associations.

## Conclusion

In this population-based cohort of adults hospitalized with AHRF, multimorbidity and socioeconomic factors were the strongest independent predictors of length of stay and hospitalization cost. Although unadjusted analyses suggested shorter length of stay and lower costs among patients admitted to hospitals with on-site AASM-accredited sleep facilities, sleep facility status was not independently associated with either outcome after adjustment. These findings suggest that accreditation status alone may not capture the inpatient care processes through which sleep medicine infrastructure could influence hospitalization outcomes. Future studies should evaluate direct measures of inpatient sleep medicine involvement and transitional care to better define their impact on acute respiratory outcomes.

## Data Availability

The data used in this study are derived from the Healthcare Cost and Utilization Project (HCUP) New York State Inpatient Database, which is subject to data-use restrictions. The analytic dataset is not publicly available. Researchers may obtain the underlying data directly from HCUP in accordance with its access policies.

